# Stasis Imaging Predicts the Risk of Cardioembolic Stroke Related to Acute Myocardial Infarction

**DOI:** 10.1101/2023.09.15.23295650

**Authors:** Elena Rodríguez-González, Pablo Martínez-Legazpi, Teresa Mombiela, Ana González-Mansilla, Antonia Delgado-Montero, Juan A. Guzmán-De-Villoria, Fernando Díaz-Otero, Raquel Prieto-Arévalo, Miriam Juárez, Maria del Carmen García del Rey, Pilar Fernández-García, Oscar Flores, Andrea Postigo, Raquel Yotti, Manuel García-Villalba, Francisco Fernández-Avilés, Juan C del Álamo, Javier Bermejo

**Affiliations:** Department of Cardiology, Hospital General Universitario Gregorio Marañón, Facultad de Medicina, Universidad Complutense de Madrid, Instituto de Investigación Sanitaria Gregorio Marañón, and CIBERCV, Madrid, Spain; Department of Mathematical Physics and Fluids, Facultad de Ciencias, Universidad Nacional de Educación a Distancia, UNED, and CIBERCV, Madrid, Spain; Department of Radiology, Hospital General Universitario Gregorio Marañón, Instituto de Investigación Sanitaria Gregorio Marañón, and CIBERSAM Madrid, Spain; Department of Neurology, Hospital General Universitario Gregorio Marañón, Instituto de Investigación Sanitaria Gregorio Marañón, Madrid, Spain; Department of Aerospace Engineering, Universidad Carlos III de Madrid, Spain; TU Wien, Institute of Fluid Mechanics and Heat Transfer, Vienna 1060, Austria; Mechanical Engineering Department; Center for Cardiovascular Biology; Institute for Stem Cell and Regenerative Medicine, University of Washington, Seattle, WA, USA

**Keywords:** ischemic stroke, ST-elevation myocardial infarction, echocardiography, blood stasis, cardioembolism

## Abstract

**Background:** In the setting of ST-segment elevation myocardial infarction (STEMI), imaging-based biomarkers could be useful for guiding oral anticoagulation for primary prevention of stroke.

**Objectives:** To test the efficacy of intraventricular blood stasis imaging for predicting a composite primary endpoint of cardioembolic risk during the first 6 months after STEMI.

**Methods:** The Imaging Silent Brain Infarct in Acute Myocardial Infarction (ISBITAMI, NCT02917213) was a prospective clinical study including patients with a first STEMI, an EF ≤ 45% and without atrial fibrillation. Patients underwent ultrasound-based stasis imaging at enrollment followed by heart and brain magnetic resonance at 1-week and at 6-month visits. From the stasis maps, we calculated the average residence time, *R*_*T*_, of blood inside the LV and assessed its performance to predict the primary endpoint. Apical longitudinal strain was quantified by speckle tracking.

**Results:** A total of 68 patients were univocally assigned to the primary endpoint. Of them, 19 patients suffered one or more events: 3 strokes, 5 silent brain infarctions, and 15 mural thromboses. No systemic embolisms were observed. *R*_*T*_ (OR: 3.28, 95% CI: 1.61-6.67, p=0.001) and apical strain (OR: 1.48, 95% CI: 1.14-1.92, p= 0.002) showed complementary prognostic value. The bivariate model showed a c-index= 0.84 (0.73-0.95) a negative predictive value of 1.00 (0.93-1.00) and positive predictive value of 0.45 (0.39 - 0.80). Results were confirmed in a multiple imputation sensitivity analysis. Conventional ultrasound-based metrics were of limited predictive value.

**Conclusions:** In patients with STEMI and LV systolic dysfunction in sinus rhythm, the risk of cardioembolic stroke can be accurately predicted by echocardiography combining stasis and strain imaging.

## Introduction

In the month following an ST-segment elevation myocardial infarction (STEMI), the adjusted risk for ischemic stroke is 30 times higher than in the general population.^1^ Oral anticoagulation is highly effective to prevent stroke, but its benefits are neutralized by its increased bleeding risk.^2-4^ Thus, preventing cardioembolic stroke in the first weeks after STEMI would benefit from a personalized risk assessment to select candidates for oral anticoagulation.

Besides atrial fibrillation (AF), two major risk factors for cardioembolism in the setting of STEMI are blood stagnation inside the left ventricle (LV) and endocardial damage.^5,6^ The latter is closely related to abnormal regional function of the ischemic territories, and therefore can be adequately addressed by strain imaging. Concerning the former, we have recently implemented a method to visualize and quantify blood stasis inside the heart from conventional echocardiographic data.^7,8^ Using this method, we have proposed a global biomarker to account for the cardioembolic risk related to blood stasis: the *residence time* (*R*_*T*_) of blood in the LV. This metric can be interpreted as the number of cycles blood spends during its transit through the LV.^8,9^ Spatiotemporal maps of the *R*_*T*_ are a valuable tool for assessing the regions prone to blood stagnation, and preliminary proof-of-concept animal and clinical studies have shown the potential of *R*_*T*_ to account for the risk of STEMI-related mural thrombosis and cerebral micro embolisms.^10,11^ Therefore, we designed the first clinical trial to prospectively assess the efficacy of stasis imaging to predict brain and heart cardioembolic events during the 6 months following STEMI. A composite primary endpoint integrated neurological events and subclinical outcomes from heart and brain imaging examinations.

## Methods

### Overall Design

For the Imaging Silent Brain Infarct in Acute Myocardial Infarction (ISBITAMI) prospective clinical study (NCT 02917213), we screened all patients admitted to our institution for the following inclusion criteria: a first STEMI with or without undergoing revascularization and an LV EF ≤ 45% at admission, age ≥ 18 years old, presence of sinus rhythm, and no history of AF. Exclusion criteria were any medical history of stroke or transient ischemic attack, ongoing oral anticoagulation treatment or a formal indication for it, any contraindication for magnetic resonance (MR) examination, a history of cardiogenic shock, history of recovered sudden death or any other potential cause of acute brain damage due to hypoperfusion, a primary valve disease ≥ 3+ severity, a diagnosis of carotid artery disease with a > 50% stenosis, history of prothrombotic disease, and reluctance to sign the written informed consent. Any history of AF (either clinical or sub clinically detected in 24-hour Holter tests or any cardiac monitoring device) was also an exclusion criterion.

Each patient underwent four imaging examinations: at screening (inclusion criteria accomplishment), in the following 24-72 hours after admission (enrollment study, echocardiography and stasis imaging), at 1 week (echocardiography and brain and heart MR), and 6 months after enrollment (echocardiography and brain and heart MR).

The Ethics Committee of the Hospital Gregorio Marañón approved the study and all patients provided written informed consent. The study was academically funded, and the Fundación para la Investigación Biomédica Hospital Gregorio Marañón was the unique sponsor. The work conforms to the principles outlined in the Declaration of Helsinki.

### Study Endpoint

The primary composite endpoint integrated the incidence of any of the following events between the inclusion and the 6-month follow-up visits: 1) a stroke or transient ischemic attack, 2) a clinically apparent peripheral systemic embolism in any arterial territory, 3) an acute or subacute silent brain infarct (SBI) lesion dated after symptom onset, assessed by brain MR, or 4) a diagnosis of LV mural thrombosis (LVT), either by contrast-echocardiography or late-gadolinium enhanced cardiac MR (CMR) studies. Patients with any of the above events between the enrollment and the 6-month follow-up visit were categorized as endpoint-positive and followed up to ensure event resolution using standard care practice. Stroke was clinically assessed, and additional imaging test (e.g. brain CT) were carried out when necessary.

### Ancillary Variables

Clinical and outcome data were collected at enrollment and 6-month visits. These included all clinical events, ongoing medications, as well as hematological and biochemical assessments and Beck Depression Inventory and Mini-mental State Examinations. We performed transcranial Doppler and carotid duplex ultrasound examinations in those patients who reached the primary endpoint to rule-out alternative extracardiac cause of embolisms. Implantable cardiac loop monitoring devices (iLINQ, Medtronic) were placed in a random sample of 31 patients to discard asymptomatic AF along the 6-month period of the study. Additional screening for AF using conventional 24-hour Holter monitoring was planned if patients reported palpitations, syncope, or any other symptom potentially caused by AF.

### Cardiac Imaging

Patients underwent four echocardiographic examinations: at screening (inclusion criteria fulfillment), in the following 24-72 hours after admission (enrollment study), at 1 week, and at 6 months. Examinations were performed using a Vivid 7 scanner and a phase-array 2-4 MHz transducer (GE Healthcare). We obtained 3-dimensional apical sequences to measure LV volumes and ejection fraction (EF) and contrast-echo sequences (intravenous Sonovue, Bracco Imaging, 1-2 mL studied using pulse inversion and a mechanical index < 0.3) to rule-out intraventricular thrombosis. Two level-3 echocardiography experts blindly analyzed the presence/absence of LVT as well as regional wall motion. Longitudinal myocardial strain was measured from 2-, 3- and 4-chamber long-axis B-mode sequences and averaged using the 16-segment model (EchoPac version 204, GE Healthcare). Apical strain was calculated as the mean of the apical segments. The E-wave propagation index (EPI) was calculated as the ratio between the E-wave velocity time integral and the LV long-axis length.^12^ Cardiac MR imaging was performed twice, at 1-week, and at 6-months after enrollment. The imaging protocol included a cine steady-state free precession imaging of LV function (SENSE X 2, repetition time: 2.4 ms, echo time: 1.2 ms, average in-plane spatial resolution: 1.6 × 2 mm, 30 phases per cycle, 8-mm slice thickness without gap) and late enhancement imaging (3D inversion-recovery turbo gradient echo sequence, pre-pulsed delay optimized for maximal myocardial signal suppression; 5-mm actual slice thickness, inversion time: 200-300 and 600 ms). Images were obtained in short axis (10 to 14 contiguous slices) and 4-, 2-, and 3-chamber views. Early and late enhancement sequences were obtained 3 to 10 min after injection of 0.1 mmol/kg of gadobenate dimeglumine (ProHance, Bracco Imaging) to assess for mural thrombosis^13^ and quantify infarct size.

### Brain Imaging

Brain MR was performed twice (at 1-week, and at 6-months visits), and it included sagittal T1-weighted images, axial diffusion weighted images, coronal T2-weigthed turbo spin echo, and axial FLAIR-T2-weigthed images. Acute or subacute diffusion weighted brain infarcts were adjudicated to the primary endpoint whenever focal lesions were greater than 3 mm hyperintense on diffusion weighted images with low apparent coefficient diffusion or pseudo normalization values were identified. Chronic ischemic injuries (not adjudicated to the primary endpoint) were diagnosed if the lesion was as intense as the cerebrospinal fluid on T2, FLAIR weighted images appeared surrounded by an hyperintense lineal rim of gliosis and showed high apparent coefficient diffusion values. Only those lesions that were unequivocally dated after AMI onset were adjudicated to the primary endpoint. All studies were interpreted by expert neuroradiologists who also differentiated these types of lesions from dilated perivascular spaces, based on their distribution and morphology.

### Intraventricular Stasis Imaging

From the echocardiograms performed at enrollment, we calculated two-dimensional, time-resolved (2D+t) blood flow fields inside the LV using color-Doppler velocimetry (echo-CDV).^14^ For this purpose, we obtained LV color-Doppler acquisitions followed by 2D cine-loops at a frame rate > 60 Hz. By imposing mass conservation, echo-CDV provides the crossbeam flow velocity, under the hypothesis of planar flow with high temporal and spatial resolutions (∼0.5 mm and 5 ms). We used these 2D+t flow fields to integrate a forced advection equation with the purpose of mapping and quantifying the residence time, *R*_*T*_. Residence time accounts for the number of cardiac cycles a volume of blood spends inside the LV.^7^ The *R*_*T*_ is computed for each “blood particle” inside the ventricle and its trajectory is followed for 8 cardiac cycles (the estimated period taken for a full washout in a normal LV).^9^ We designed a color-coded scale to generate video maps of RT of blood in the LV. In each frame, the color scale represents the number of cardiac cycles a blood particle has spent inside the chamber; dark blue represents “fresh” blood recently entering the LV, whereas dark red represent stasis regions in which with blood is retained for at least 4 cardiac cycles (**see Supplementary Material** and **Videos** for details). At the end of the 8^th^ beat, we collected the average *R*_*T*_ inside the LV as a single scalar. The reproducibility of stasis metrics has been reported elsewhere.^9^ Additional methodological details are summarized in the **Supplemental Material & Figure S1**.

### Statistical Analysis

Sample size of the study was established in n= 92 patients to achieve an 85% power for achieving significance (p< 0.05) with a c-index> 0.75, assuming a 15% incidence of the primary endpoint and a 15% attrition rate. Data are described as median [interquartile range] except otherwise indicated. We used Wilcoxon rank-sum and Fisher exact tests to compare quantitative variables and proportions, respectively. Odds-ratios and their 95% confidence intervals (CIs) are reported for these models. The interplay between clinical predictors, *R*_*T*_, and the primary endpoint was further investigated using Pearson correlation analyses. We used ROC analyses to estimate the c-index, its 95% CI, and its statistical significance. Cutoffs were selected by the Youden’s method, weighted to penalize for false negatives. Medians (and 95% CIs) of performance metrics of these cutoffs (sensitivity, specificity, positive and negative predictive values) were calculated by bootstrap of 2,000 replicates.

The analyses are reported for patients who reached the end of the follow-up protocol or could be allocated to the primary endpoint because they had an event before (**Figure 1**). We performed a sensitivity analysis to avoid verification bias.^15^ For that purpose, we used multiple imputation by chained equations of the missing values of the individual events that constitute the primary endpoint in the patients with stasis imaging lost for follow-up. These 1,000 imputed datasets were used for analyses of the primary endpoint, and median metrics of diagnostic performance and their 95% CIs were calculated using Rubin’s rule. By design, the endpoint time-window was defined between inclusion and the 6-month follow-up visit. However, as 2 patients showed LVT in the enrollment study, we also evaluated stasis performance to predict embolism after this time point (removing these 2 patients from the analyses). Statistical analyses were performed in R (v. 4.1.3) and p-values < 0.05 were considered significant.

**FIGURE 1:**
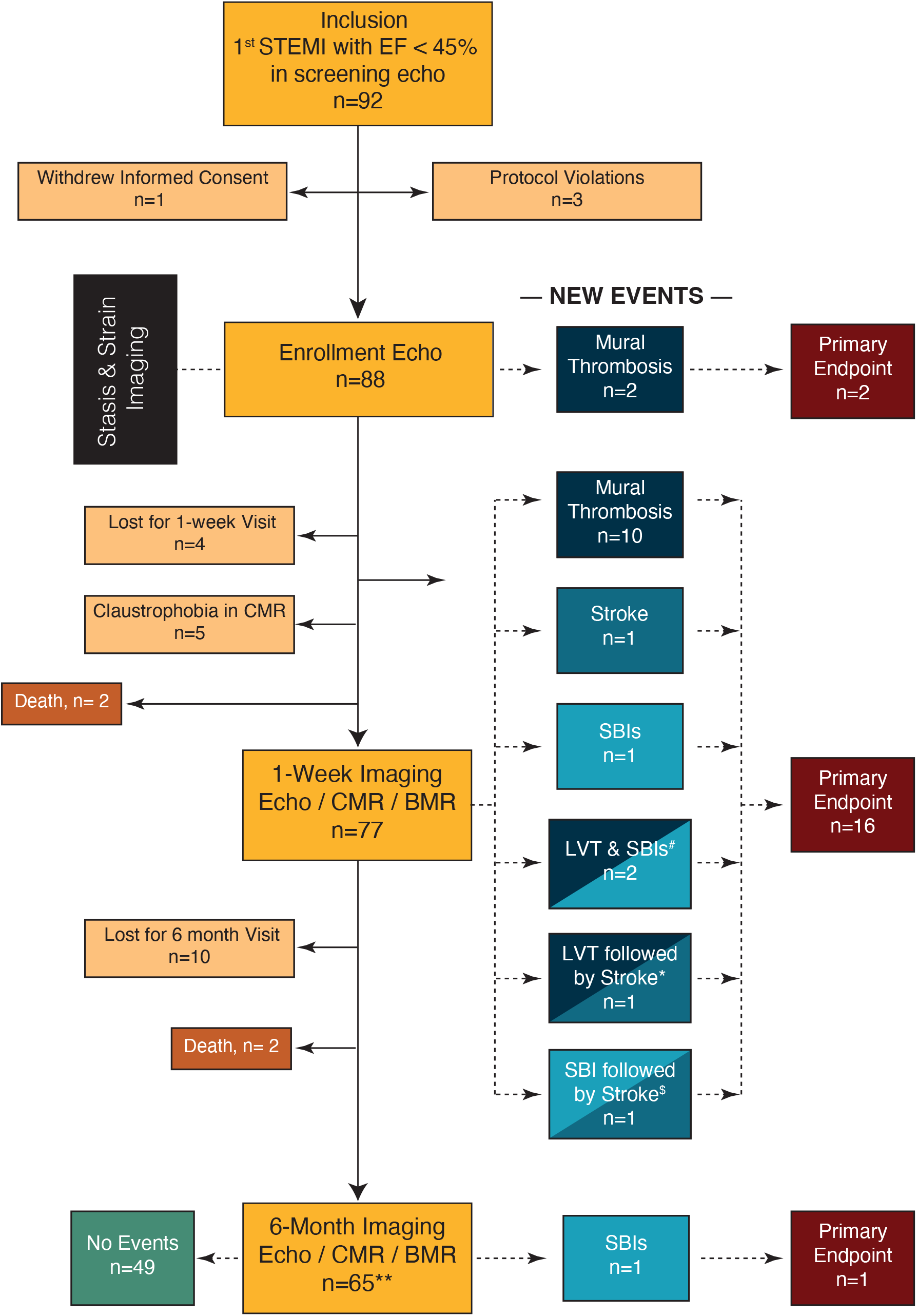
Flow Diagram of the ISBITAMI study and the Primary Endpoints. CMR/BMR: Cardiac & Brain Magnetic Resonance. LVT: Mural Thrombosis. SBI: Silent Brain Infarction. ^#^: Two patients simultaneously showed SBIs and LVT. *: One patient suffered a stroke 4 days after LVT was imaged despite being under OAC therapy. ^$^: One patient suffered a stroke 1 week after a SBI was imaged. **: Twelve patients were lost during follow-up. Of them, 3 patients had already reached the endpoint, recorded on the 1-week visit.

## Results

### Study Population

Patient recruitment started in January 2017 and last patient follow-up ended in January 2022. From an initial group of 92 screened patients, a total of 68 patients were univocally assigned to the primary endpoint: 65 patients completed the six-month follow-up and 3 patients without follow-up but who reached positive endpoint in the 1-week visit (**Figure 1)**.

Median [IQR] age was 58 [51-67] years old, and 22% patients were women (**Table 1**). Sixty-four patients underwent primary percutaneous coronary intervention, and STEMI location was anterior in 61. Four patients did not undergo primary PCI either because they underwent a PCI following thrombolysis (n= 2) or because a conservatory strategy was chosen due to advance age (n= 1) or PCI disesteemed due to unsuitable anatomy (n= 1). Ejection fraction by echocardiography at enrollment was 41 [35-47]%, and values obtained by CMR at 1-week were similar (**Table 2**).

**Table 1.**
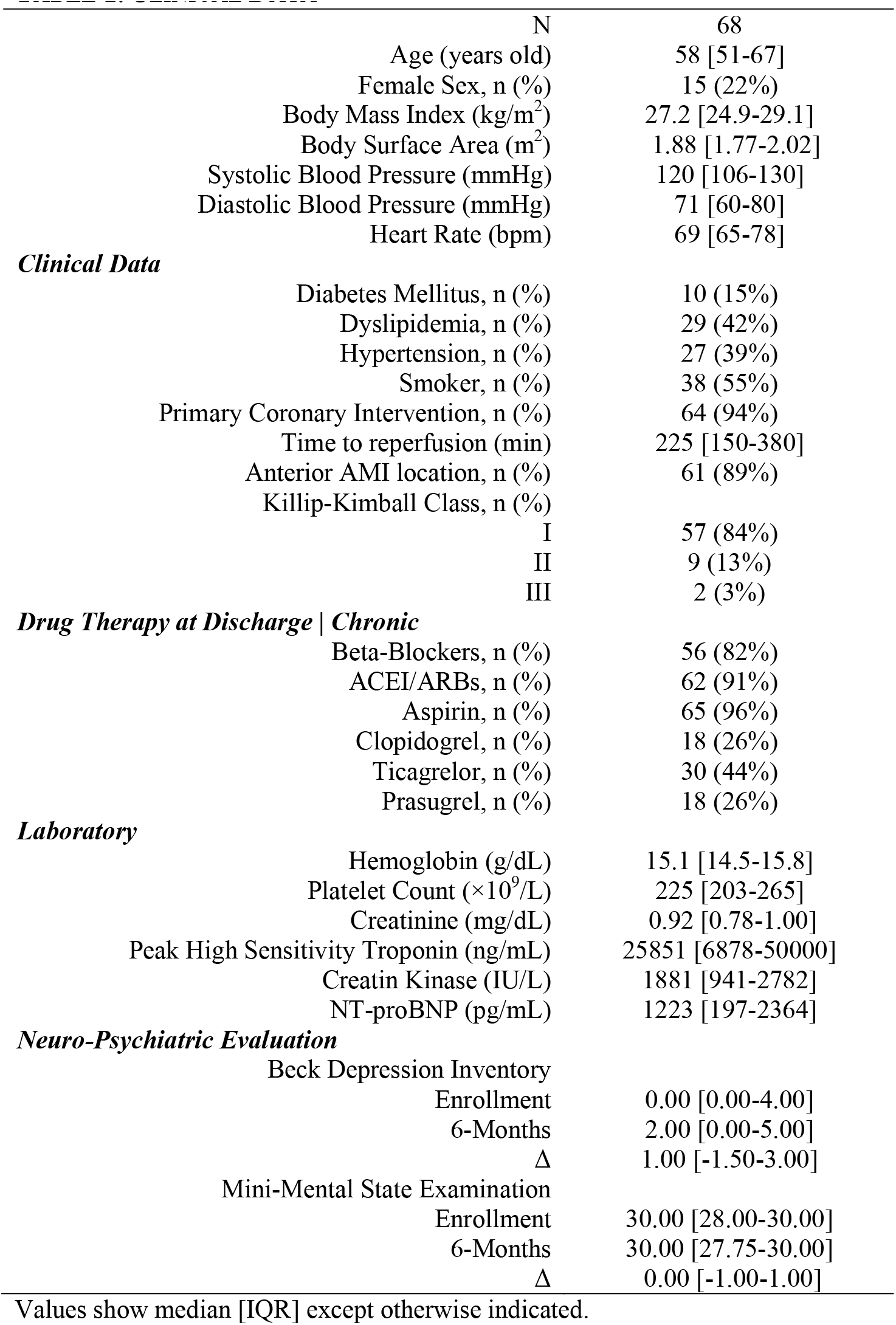
Clinical Data.

**Table 2:** Cardiac Imaging.

|  | Imaging Time<br>N | Enrollment<br>68 | 1-Week Visit<br>68 | 6-Month Visit<br>65 |
| --- | --- | --- | --- | --- |
| **Echocardiography** |  |  |  |  |
| LV End-Diastolic Volume Index (mL/m<sup>2</sup>) |  | 49 [43-61] | 50 [43-60] | 58 [47-67] |
| LV End-Systolic Volume Index (mL/m<sup>2</sup>) |  | 29 [24-36] | 30 [22-34] | 29 [23-35] |
| Stroke Volume Index (B mode) (mL/m<sup>2</sup>) |  | 20 [18-24] | 23 [19-26] | 27 [24-32] |
| Stroke Volume Index (Doppler) (mL/m<sup>2</sup>) |  | 26 [21-28] | 27 [22-32] | 30 [26-36] |
| LV Ejection Fraction (%) |  | 41 [35-47] | 43 [38-51] | 50 [43-57] |
| Sphericity Ratio |  | 0.53 [0.48-0.58] | 0.54 [0.50-0.58] | 0.57 [0.51-0.60] |
| E-wave velocity (cm/s) |  | 0.53 [0.45-0.66] | 0.60 [0.49-0.71] | 0.61 [0.48-0.77] |
| A-wave velocity (cm/s) |  | 0.68 [0.57-0.76] | 0.67 [0.56-0.75] | 0.63 [0.51-0.73] |
| E/A wave ratio |  | 0.78 [0.63-1.11] | 0.81 [0.68-1.28] | 0.96 [0.73-1.50] |
| E/e' |  | 8.9 [7.2-10.4] | 9.41 [8.02-10.88] | 8.3 [7.0-9.8] |
| E-wave penetration index |  | 0.90 [0.74-1.12] | 1.04 [0.82-1.28] | 1.25 [0.86-1.59] |
| Global Peak Systolic Longitudinal Strain (%) |  | -9.2 [-10.9- -8.1] |  |  |
| Apical Peak Systolic Longitudinal Strain (%) |  | -7.3 [-8.8- -4.7] |  |  |
| Wall Motion Score Index |  | 2.00 [1.81-2.13] |  |  |
| **Stasis Imaging** |  |  |  |  |
| Intraventricular Blood Residence Time (cycles) |  | 2.94 [2.54-3.57] |  |  |
| **CMR** |  |  |  |  |
| LV End-Diastolic Volume Index (mL/m<sup>2</sup>) |  |  | 87 [81-101] | 94 [83-108] |
| LV End-Systolic Volume Index (mL/m<sup>2</sup>) |  |  | 50 [44-60] | 49 [40-63] |
| Stroke Volume Index (mL/m<sup>2</sup>) |  |  | 37 [33-43] | 43 [39-49] |
| LV Ejection Fraction (%) |  |  | 41 [38-47] | 48 [40-53] |
| LV Mass Index (g/m<sup>2</sup>) |  |  | 60 [52-64] | 52 [44-56] |
| Infarct Size (g) |  |  | 25 [18-38] | 20 [14-29] |
| Infarct Size (%) |  |  | 0.27 [0.16-0.33] | 0.21 [0.14-0.28] |

Values show median [IQR] except otherwise indicated.

### Cardioembolic Events

After the 6-months follow up period, 23 events were recorded in 19 of the 68 studied patients (28%). Three patients suffered an ischemic stroke. Of them, one occurred after the diagnosis of LVT despite having started OAC treatment. Another stroke took place one week after finding an SBI. Fourteen patients showed LVT, two of them showing accompanying SBIs in the same 1-week visit. Finally, two patients were diagnosed of SBIs without additional events (one in the 1-week visit and the other one in the 6-month visit). No peripheral systemic embolisms were clinically identified (**Table 3**).

**Table 3:** Composite Endpoint.

|  | N (%) | Comments |
| --- | --- | --- |
| **Total Patients** | 68 |  |
| **Patients who reached Composite Endpoint** | 19 (28%) |  |
| @ Enrollment (N= 68) | 2 (3%) | 2 LVT. |
| @ 1-week visit (N= 68) | 16 (23%) | 3 Stroke (1 coexisting with a SBI and 1 coexisting with LVT). 12 LVT (2 coexisting with SBIs); 1 SBI; |
| @ 6-month visit (N= 65) | 1 (1%) | 1 SBI. |
| **Left Ventricular Mural Thrombosis** |  |  |
| @ Enrollment (N= 68) | 2 (3%) | Not present @ screening echo. Both disappeared after 1-week OAC and were not present @ 1-week CMR. |
| @ 1-week CMR (N= 68) | 13 (19%) | 11 disappeared after 6-month OAC and were not present @ 6-month CMR. 1 was followed by a stroke 4-days after the visit despite OAC. |
| @ 6-month CMR (N= 65) | 0 (0%) | 2 still present from 1-week CMR. |
| Total (N= 68) | 15 (22%) |  |
| **Silent Brain Infarct** |  |  |
| @ 1-week Brain MR (n= 68) | 4 (6%) | 2 coexisting with LVT @ 1-week CMR. |
| @ 6-month Brain MR (n= 65) | 1 (2%) | 1 not present @ 1-week BMR and without LVT. |
| Total (N= 68) | 5 (7%) |  |
| **Stroke** |  |  |
| Total (N= 68) | 3 (4%) | 1<sup>st</sup> 4-days after LV mural thrombosis @ 1-week CMR and under OAC; 2<sup>nd</sup> one week after SBI found @ 1-week BMR; 3<sup>rd</sup> without LV mural thrombosis or SBI in the 1-week MRI. |
| **Peripheral Embolism (other territories)** |  |  |
| @ Enrollment (N= 68) | 0 (0%) |  |
| @ 6-month Follow up (n= 65) | 0 (0%) |  |

OAC: oral anticoagulation therapy; LVT: LV mural thrombosis. SBI: silent brain infarct. CMR: cardiac magnetic resonance; BMR: brain magnetic resonance.

Carotid duplex and transcranial Doppler ruled out alternative etiologies of the neurological events in all patients with SBI or stroke. No AF was identified during the 6-month period in the 31 randomly selected subjects who received an implantable cardiac rhythm monitoring device. There were no differences in clinical, hematological, or biochemical variables among patients with and without the primary endpoint. There were no differences in Beck Depression Inventory or Mini-Mental State Examinations amongst patients with and without the primary endpoint, neither at enrollment nor at the 6-month follow-up visit. Myocardial infarctions were of an anterior location in all patients with a primary endpoint.

### Imaging Predictors of the Primary Endpoint

Patients with a primary endpoint showed larger myocardial infarct size by CMR than those patients without endpoint (32 [29-37]% vs. 21 [15-31]% of total myocardial mass, respectively p= 0.04). Enrollment echocardiographic examinations showed lower values of EF in patients with a primary endpoint (38 [34-43]%) than in those without endpoint (42 [36-51]%, p= 0.02). Also, apical strain was higher in patients with a primary endpoint than in those free of events: -4.7 [-7.2- -3.6] vs -7.9 [-10.1- -6.0]% (p< 0.01).

The c-index of EF to predict the primary endpoint was 0.68 (95% CI= 0.55-0. 81). However, this predictive power was related to the fact that EF was < 50% in all patients with a primary endpoint; the primary endpoint was not identified in any patient whose EF reached 50% from inclusion to enrollment. Among patients who persisted with an EF < 50%, the c-index of EF was 0.55 (0.38-0.71, p= 0.6). The c-index of apical strain to predict the primary endpoint was 0.75 (0.64-0.88) with an odds-ratio of 1.48 (95% CI: 1.14-1.92, p= 0.002, **Table 4)** per unit. The c-index of *R*_*T*_ was 0.79 (0.68-0.90) with an OR of 3.28 (1.61-6.67) per cycle (p= 0.001, **Table 4** and **Figure 2**). When excluding the patients with events found in the enrollment (2 LVT, **Figure 1**), the c-index of *R*_*T*_ was 0.82 (0.71-0.92) with an OR of 3.73 (1.75-7.97). Correlation of *R*_*T*_ with EF and infarct size (by CMR) was R= -0.30 and 0.22 (p=0.005 and 0.07), respectively.

**Table 4:** Performance of Stasis and Strain Imaging To Predict the Primary Endpoint.

| Index | c-index | Threshold | Sensitivity | Specificity | PPV | NPV |
| --- | --- | --- | --- | --- | --- | --- |
| Residence Time | 0.79<br>(0.68-0.90)* | > 2.75<br>cycles | 0.95<br>(0.84-1.00) | 0.44<br>(0.28-0.59) | 0.41<br>(0.34-0.48) | 0.95<br>(0.88-1.00) |
| Apical Strain | 0.75<br>(0.64-0.88)* | -8.1 % | 0.94<br>(0.83-1.00) | 0.44<br>(0.29-0.61) | 0.41<br>(0.35-0.50) | 0.95<br>(0.85-1.00) |
| Residence Time + Apical Strain | 0.84<br>(0.73-0.95)* | 0.14 | 1.00<br>(0.84-1.00) | 0.52<br>(0.35-0.93) | 0.45<br>(0.39 -0.80) | 1.00<br>(0.93-1.00) |

\*p< 0.05; thresholds selected by Youden's criterion weighting sensitivity. Values are shown for bootstrap values of 2,000 replicates. PPV: Positive Predictive Value. NPV: Negative Predictive Value.

**FIGURE 2:**
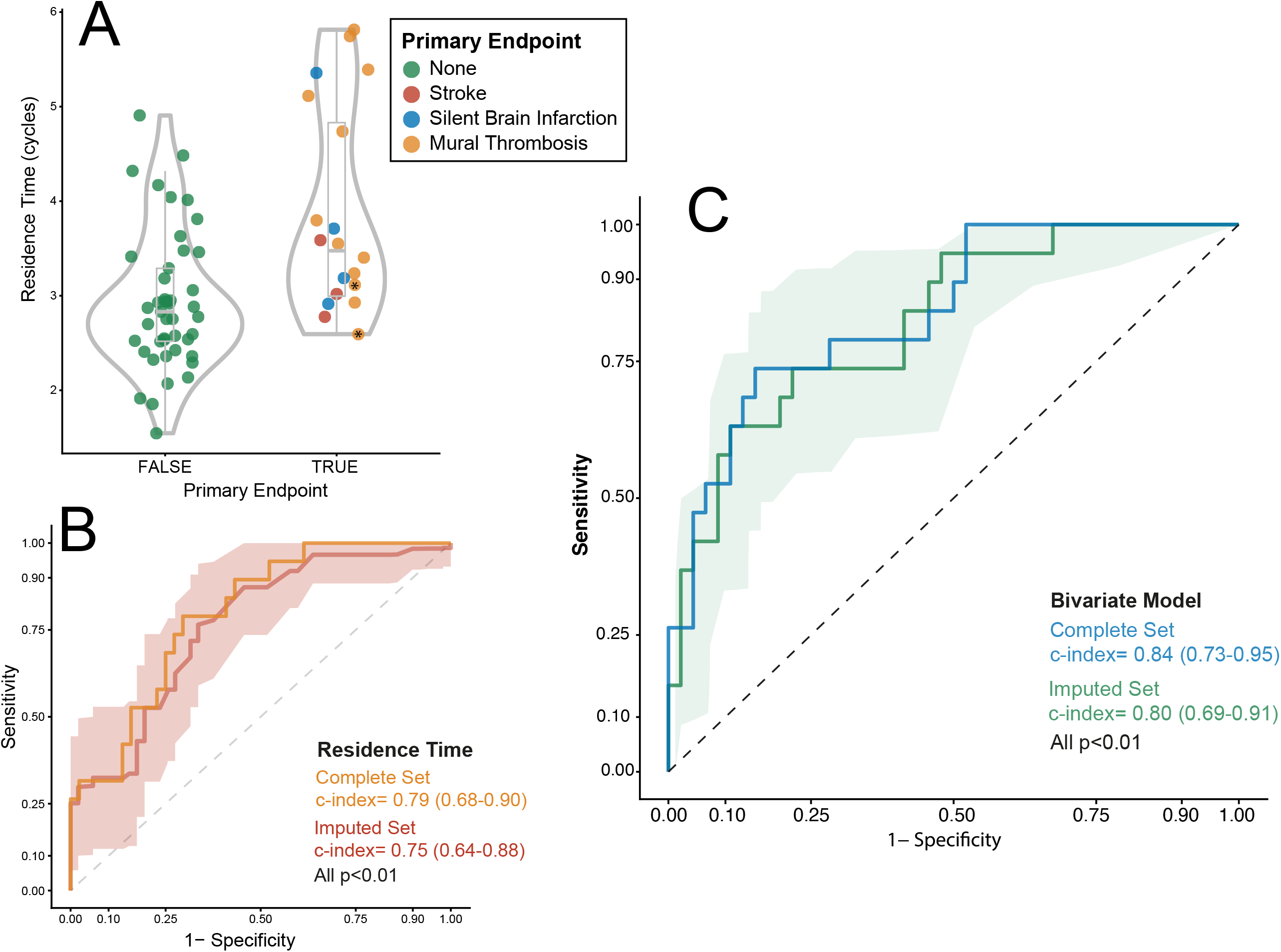
Relationship between Residence Time and the Primary Endpoint. **Panel A:** Violin and boxplots; the primary endpoint is colored upon its etiology. *: Residence time of patients with mural thrombosis found at enrollment. **Panel B:** ROC curve for the performance of *R*_*T*_ for predicting the primary endpoint (complete and imputed sets**). Panel C:** ROC curve for the performance of the bivariate model (*R*_*T*_ + apical strain) for predicting the primary endpoint (complete and imputed sets). The ribbons on Panels B & C show the sensitivity and 95% confidence interval of the ROC-curve calculated for the imputed set.

Apical strain correlated neither with *R*_*T*_ (R= 0.17, p= 0.07) nor infarct size (R= 0.21, p= 0.08). Predictive accuracy was highest in the bivariate model including both variables: c-index= 0.84 (0.73-0.95), both apical strain (OR= 1.42, 1.07-1.88) and *R*_*T*_ (OR= 3.02, 1.36-6.72) retaining significant predictive value (p= 0.01 and 0.006, respectively, **Figure 2**). Sensitivity, specificity, positive and negative predictive values of the bivariate model were1.00 (0.84-1.00), 0.52 (0.35-0.93), 0.45 (0.39-0.80) and 1.00 (0.93-1.00), respectively. The c-index of the bivariate model was 0.74 (0.60-0.88) amongst patients with an EF< 50%. The final model for the risk of the primary endpoint was log(risk)= -2.6 + 0.35?strain (%) + 1.1?*R*_*T*_ (cycles). C-index of the bivariate model was similar, 0.73 (0.59-0.88), amongst those patients with an EF< 50% and with event occurrence after enrollment (17 patients with events, **Figure 1**). The sensitivity analysis confirmed similar performance of apical strain and *R*_*T*_ to predict the primary endpoint in the imputed set (**Table 1S**). **Figure 3** and **Videos S1-S2** show illustrative examples from patients with and without the primary endpoint. **Video S3** show the *R*_*T*_ distribution in a healthy volunteer for comparison.

**FIGURE 3:**
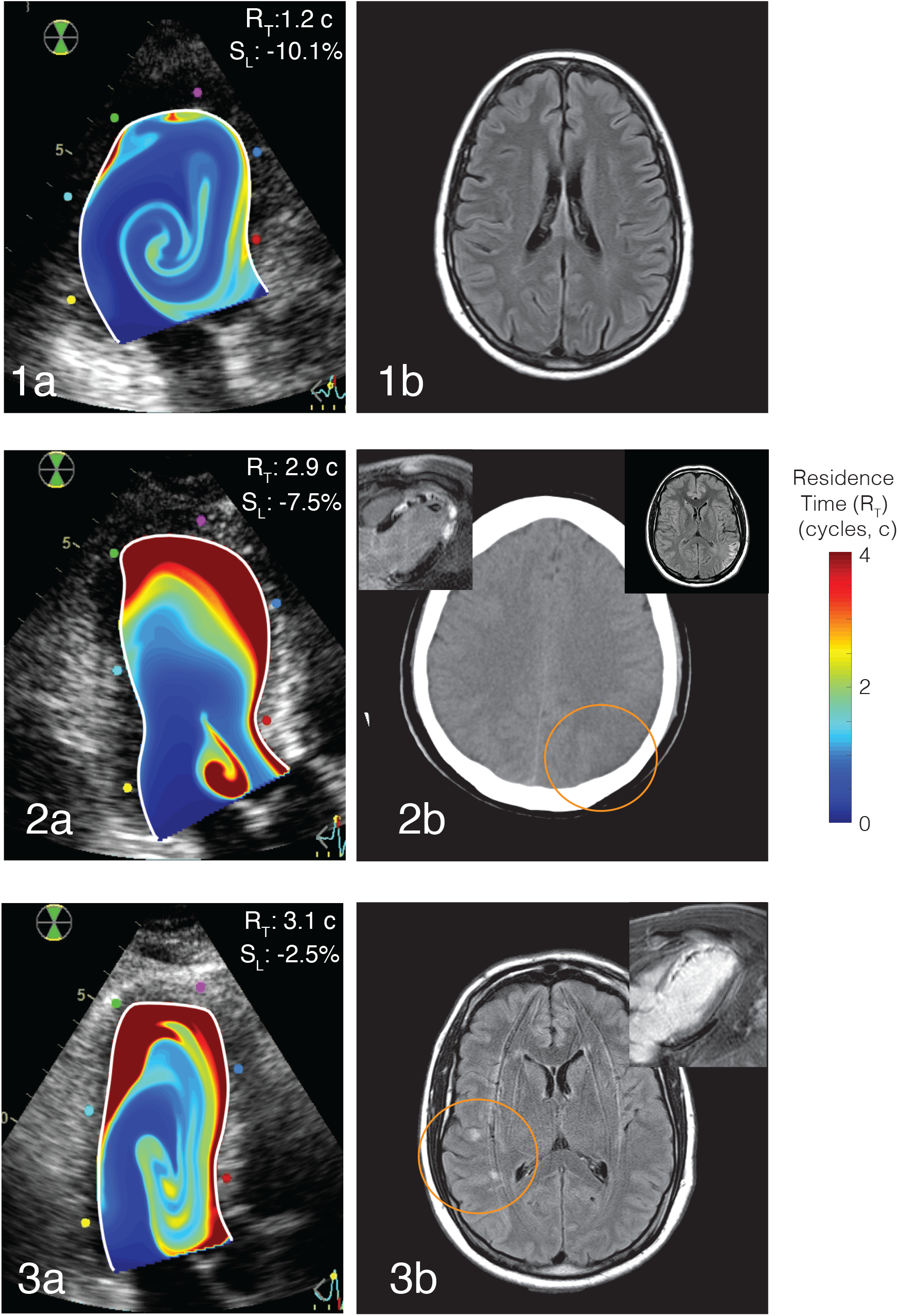
Representative Examples of Residence Time Imaging and the Primary Endpoint. **Panel 1 (a-b)**: *R*_*T*_ mapping and brain MR of a patient with no primary endpoint. **Panel 2 (a-b)**: *R*_*T*_ mapping and conventional imaging (brain computed tomography and inserts of CMR and brain MR) of a patient with an ischemic stroke and LV mural thrombosis. **Panel 3 (a-b):** *R*_*T*_ mapping and brain imaging and insert of CMR of a patient with a SBI. *R*_*T*_ mapping is overlaid on the B-mode echocardiogram. Upper-right corner in **Panels 1-a, 2-a & 3-a**: individual values of apical strain, *S*_*L*_, and residence time, *R*_*T*_.

## Discussion

The ISBITAMI is a proof-of-concept study demonstrating the value of combining strain imaging and stasis biomarkers to account for cardioembolic risk after STEMI. By exploiting the well-known mechanistic associations between stasis, local damage, and thrombosis, we implemented an ultrasound-based model that accurately predicted the incidence of cardioembolic events in patients with STEMI. The prediction efficacy suggests a potential for grounding anticoagulation therapy on strain and stasis imaging (**Figure 4: Central Illustration**).

**FIGURE 4:**
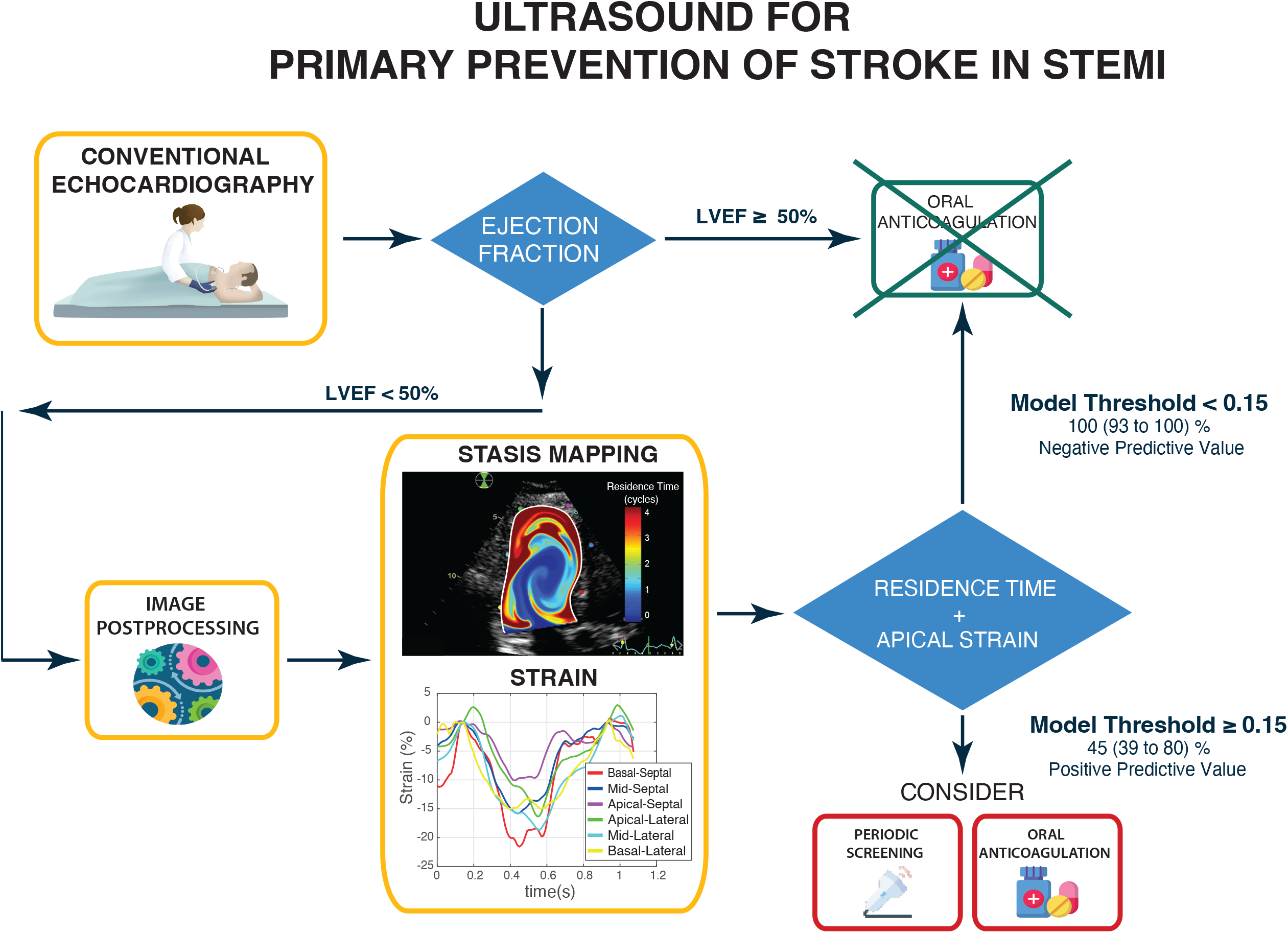
CENTRAL ILLUSTRATION: Potential Algorithm for Indicating Anticoagulation Based on the Results of Stasis Imaging and Strain

To our knowledge, this is the first study prospectively addressing SBIs in the setting of STEMI. SBIs designate those lesions of brain parenchyma that meet the imaging characteristics of an infarct, but which are not associated with clinical signs or symptoms of stroke or transient ischemic attack. SBIs are frequently identified in patients with systolic dysfunction,^16^ a condition leading to cognitive impairment and which doubles the risk for stroke.^17^ Brain MR is the gold standard technique for the diagnosis of SBI, and diffusion-weighted images allow dating SBI lesions within a few hours of their occurrence, therefore discriminating acute, subacute, and chronic injuries. SBIs are a sensitive proxy of cardioembolic risk in several cardiac diseases and procedures, as well as a source of disability and mortality *per se*.^18^ The results of our study prospectively demonstrate a temporal association between SBIs and STEMI, which has been indirectly suggested by the frequent finding of myocardial scars in patients with SBIs.^19^ Nevertheless, whether both indices remain predictors of cardioembolism in other settings needs to be addressed. We observed a 6-month incidence of SBIs of 6% in our STEMI population, and half of patients with SBI also showed LVT. In one case, the identification of SBI was followed by an ischemic stroke a few days later. Furthermore, one stroke took place in a patient in which LVT had been identified and anticoagulation initiated —in the setting of this comprehensive research-related imaging workup. This complication highlights the limitations of current stroke prevention guided exclusively by the visualization of mural thrombosis,^5,20^ and illustrates the potential advantage of anticipating anticoagulation before local thrombosis develops. Although SBIs can be caused by the cardiac catheter interventions *per se*,^21^ the observation of concomitant mural thrombosis in two patients and the subsequent stroke in another one suggests catheter manipulation as a highly unlikely source of SBIs in our cohort.

Blood stasis is a key factor Virchow’s triad that determines thrombosis.^6^ However, current methods for quantifying stasis in the heart are qualitative and limited. Although spontaneous contrast is related to the risk of thrombosis both in the left atrium^22^ and the LV^23^ spontaneous contrast is highly dependent on operator, equipment, and non-fluidic rheological factors.^24^ Technological advances in the last decade open the opportunity for obtaining time-resolved two- or even three-dimensional flow velocity fields from imaging modalities.^7,25^ Applying the physical laws of fluid dynamics to these velocity fields, it is possible to derive quantitative indices that account for the transport of blood inside the chambers, the interaction between incoming fresh blood and the remanent pool, and the residual stasis. We have demonstrated that the average *R*_*T*_ of blood in the LV closely correlates with the number of high-intensity signals detected by carotid Doppler ultrasound in a porcine model of STEMI.^11^ In another pilot clinical study, *R*_*T*_ was also higher in patients who showed mural thrombosis during the subacute phase of STEMI.^10^ Blood transport in the LV is a complex phenomenon, governed by fluid-structure and fluid-dynamical interplays which are, in turn sensitive to early versus late filling fractions, the degree of chamber emptying, and the development of the diastolic vortex ring.^7,9,26^ This explains why the risk of mural thrombosis after STEMI is related to and abnormal apical flow transport.^27,28^ These aspects can only be indirectly inferred by visual inspection of velocity fields but can very accurately analyzed and quantified using stasis imaging.

The second factor of Virchow’s triad is related to the tissular changes in the vessel walls. Endocardial injury and collagen exposure locally activates the coagulation system, and the extension of local damage is closely related to the risk of thrombosis. In the setting of STEMI, the degree of impairment of myocardial longitudinal strain is closely related to the transmural extent of myocardial necrosis.^29^ In our study, apical strain was related to cardioembolic risk^30^ but it was not mediated by increased intraventricular stasis,^31^ as apical strain did not correlate with *R*_*T*_.

### Clinical Implications

The ISBITAMI study opens the opportunity for a randomized clinical trial assessing the efficacy of oral anticoagulation in selected patients guided by stasis imaging. Anticipating a 45% probability of suffering a cardioembolic event based on imaging (the positive predictive value of our model) is a risk-benefit ratio that justifies initiating anticoagulation in this subgroup of patients with STEMI. Idiosyncratic coagulation blood-related factors may potentially account for the finding of only a 45% positive predictive value of the imaging model. Interestingly, these coagulation factors can be efficiently incorporated to the fluid-dynamics models to obtain integrated metrics of thrombosis.^32^ A negative predictive value > 95% suggests very few patients at risk would be left untreated. Further external validation should confirm these findings.

After an embolic stroke of unknown source, also the benefit of anticoagulation needs to be balanced against bleeding,^4^ and stasis imaging may also be useful in this setting. Remarkably, stasis can also be quantified for blood in the left atrium and the left atrial appendage.^33^ Because LV systolic dysfunction, atrial myopathy and disturbed intra-atrial flow are very prevalent in patients with embolic stroke of unknown source,^34^ stasis imaging may be also useful in this condition. Stasis imaging may also provide insight on the relationship between subclinical LV systolic dysfunction and silent cerebrovascular disease.^4^

### Limitations

Designed as a monocentric study, the number of patients and events was relatively small. Thus, results should be validated in a larger multicentric clinical trial. Despite the study inclusion criteria established an EF< 45% in the screening exam, it was > 50% in 20 patients at the time of the enrollment echocardiogram (performed within 72 hours following inclusion) due to early recovery of systolic function after revascularization. The primary endpoint integrated events of different clinical relevance. Although it may lower the prognostic implications of our findings, the composite primary endpoint is backed by 1) the well-established relationship between neurological and imaging outcomes summarized above, and 2) the poor prognostic implications of LV mural thrombosis after STEMI.^20^ SBIs and neurological events cannot be unequivocally attributed to a cardioembolic origin and alternative etiologies are plausible. To minimize this limitation, we performed comprehensive etiological workup to all patients with SBIs and implanted cardiac monitoring devices in a random sample of patients. Nevertheless, it is impossible to completely exclude small vessel disease or subclinical AF as causing the identified lesions. The role of catheterization procedures as a source of SBIs *per se* has been discussed above. The 25% incidence of the primary endpoint may seem higher than in other series. However, this could be due to the strict inclusion criteria, focusing on patients with LV systolic dysfunction at admission and undergoing comprehensive serial cardiac and brain imaging assessment. As anticipated, not all patients underwent full follow-up procedures for adjudication of the primary endpoint. However, potential verification bias was excluded by a comprehensive imputation sensitivity analysis.

Regarding stasis imaging, there are some limitations. Although our method is based on 2D transthoracic echocardiography and assumes a planar-flow distribution, appropriate validation studies show that the error related to this assumption is small, and the method shows good reproducibility.^9,35,36^ Nevertheless, whether the performance of stasis indices is improved when calculated from 3-dimensional CMR data deserves further investigation.^8^

## Conclusions

In patients with STEMI, cardioembolic risk can be predicted using bedside echocardiography combined with stasis imaging. This imaging modality may be useful for personalizing primary stroke prevention in patients with impaired systolic function and no history of AF.

## Data Availability

The data that support the findings of this study are available from the corresponding author upon reasonable request

## Acknowledgments

We are in debt with all the personnel of the Department of Cardiology of the Hospital General Universitario Gregorio Marañón and with Ana Fernández Baza for her assistance in the logistics of the ISBITAMI study.

## Supplemental Materials

Supplemental Methods

Table S1

Figures S1

Videos S1-S3

